# BIOLOGICAL RHYTHMS IN COVID-19 VACCINE EFFECTIVENESS

**DOI:** 10.1101/2022.10.31.22281719

**Authors:** Guy Hazan, Or A. Duek, Hillel Alapi, Huram Mok, Alex Ganninger, Elaine Ostendorf, Carrie Gierasch, Gabriel Chodick, David Greenberg, Jeffrey A. Haspel

## Abstract

**Importance:** Circadian rhythms affect fundamental immune processes, but how this translates to clinical outcomes like real-world vaccine effectiveness is unclear.

**Objective:** To examine associations between Coronavirus Infectious Disease 2019 (COVID-19) vaccination timing and effectiveness.

**Design, Setting, and Participants:** Retrospective cohort study of database records from Maccabi Healthcare Services (MHS), a major Israeli Health Maintenance Organization (HMO). We included all individuals over 12 with at least one timestamped vaccine dose and no documented COVID-19 infection prior to completing the initial 2-dose immunization series (n=1,515,754, 99.2% receiving BNT162b2). Database records spanned December 19, 2020, to April 25, 2022, encompassing two spikes in COVID infection dominated by the delta (B.1.617.2) and omicron (B.1.1.529) SARS-CoV-2 variants.

**Main Outcomes and Measures:** Outcomes included COVID-19 breakthrough infection and COVID-19 associated emergency department (ED) visits. Our main comparison was between patients vaccinated exclusively during morning hours (8:00-11:59), afternoon (12:00-15:59), or evening hours (16:00-19:59). We employed Cox multivariate regression to adjust for differences in age, sex, and co-morbidities.

**Results:** Breakthrough infections differed based on vaccination time, with lowest rates associated with late morning to early afternoon, and highest rates with evening vaccination. Vaccination timing remained significant after adjustment for patient age, sex, and co-morbidities (HR=0.88 afternoon vs. evening, [95% CI 0.87-0.90]). Results were consistent in patients who received the basic two-dose vaccine series and who received booster doses. The relationship between COVID-19 immunization time and breakthrough infection risk was sinusoidal, consistent with a biological rhythm in vaccine effectiveness. Vaccination timing altered breakthrough infection risk by 8.6-25% in our cohort, depending on patient age and dose number. The benefits of daytime vaccination were concentrated in younger and elderly patients. In contrast to breakthrough infections, COVID-19 related ED visits correlated with age and medical comorbidities but not with time of vaccination.

**Conclusions and Relevance:** We report a significant association between the time of COVID-19 vaccination and its clinical effectiveness in terms of breakthrough infection. These data have implications for mass vaccination programs.

**KEY POINTS:** 

**Question:** Does the time of day patients receive their COVID-19 vaccinations influence their clinical benefit?

**Findings:** In this population-level cohort study that included 1,515,754 individuals aged 12 and over, COVID-19 vaccination during the late morning to early afternoon was associated with fewer breakthrough infections compared to other times. Vaccination timing altered breakthrough infection risk by 8.6-25%, depending on patient age and dose number.

**Meaning:** Prioritizing children and the elderly for late morning to early afternoon immunization could improve the effectiveness of mass vaccinations against COVID-19, and potentially other infectious diseases.

## INTRODUCTION

Circadian rhythms are daily oscillations in biological function that enable organisms to align their physiology to the day-night cycle^1,2^. These rhythms emanate from a genetically encoded molecular clock that regulates gene expression and thereby organizes cellular functions into daily cycles^1,2^. Among the activities organized by the circadian system are basic immune processes^3-5^, including the innate inflammatory response^6-12^, cellular egress from the bone marrow^13^, leukocyte trafficking to organs and lymph nodes^14-16^, and T cell responses to antigen^17,18^. Some theorize that the circadian rhythms in humans should translate, directly or indirectly, into preferable times of day to vaccinate patients^1,2,19-21^. However, clinical studies differ on whether rhythms exist in vaccine responses, including those against severe acute respiratory syndrome coronavirus 2 (SARS-CoV-2), the cause of coronavirus infectious disease-19 (COVID-19)^22-31^. While important, prior studies had limited sample sizes and focused on markers of immunogenicity like antibody titers rather than clinical outcomes. Here, we examine how the timing of COVID-19 vaccination relates to clinical protection using large-scale observational data.

## METHODS

### Study design and setting

This retrospective cohort study analyzes database records from Maccabi Healthcare Services (MHS) in Israel. MHS is the second largest state-mandated, not-for-profit, health maintenance organization (HMO) in Israel with over 2.6 million members, constituting one-quarter of the country’s population. MHS maintains extensive medical, demographic, and anthropometric information linked to a nation-wide electronic medical record (EMR) servicing the entire Israeli civilian population. The MHS database captures all clinical encounters, diagnoses, medications, and laboratory data for its participants anywhere within the country regardless of setting, including all clinical activity and diagnostic testing related to COVID-19. All data were de-identified prior to analysis. Since this was a retrospective study, patient informed consent was not required. For identities of those who analyzed the data see “Study Oversight” in the **eAppendix1** of the **Supplement**. The MHS ethical committee approved the study.

### Study period

We analyzed data from December 19, 2020 (the first day of vaccine administration in the study population) to April 25, 2022 (the last day of data extraction). This timespan encompasses two spikes in COVID infection dominated by the delta (B.1.617.2) and omicron (B.1.1.529) SARS-CoV-2 variants (**eFigure 1** in the **Supplement**, black and blue bars). For background information on the COVID-19 vaccine rollout in Israel see **eAppendix2** in the **Supplement**.

**Figure 1.**
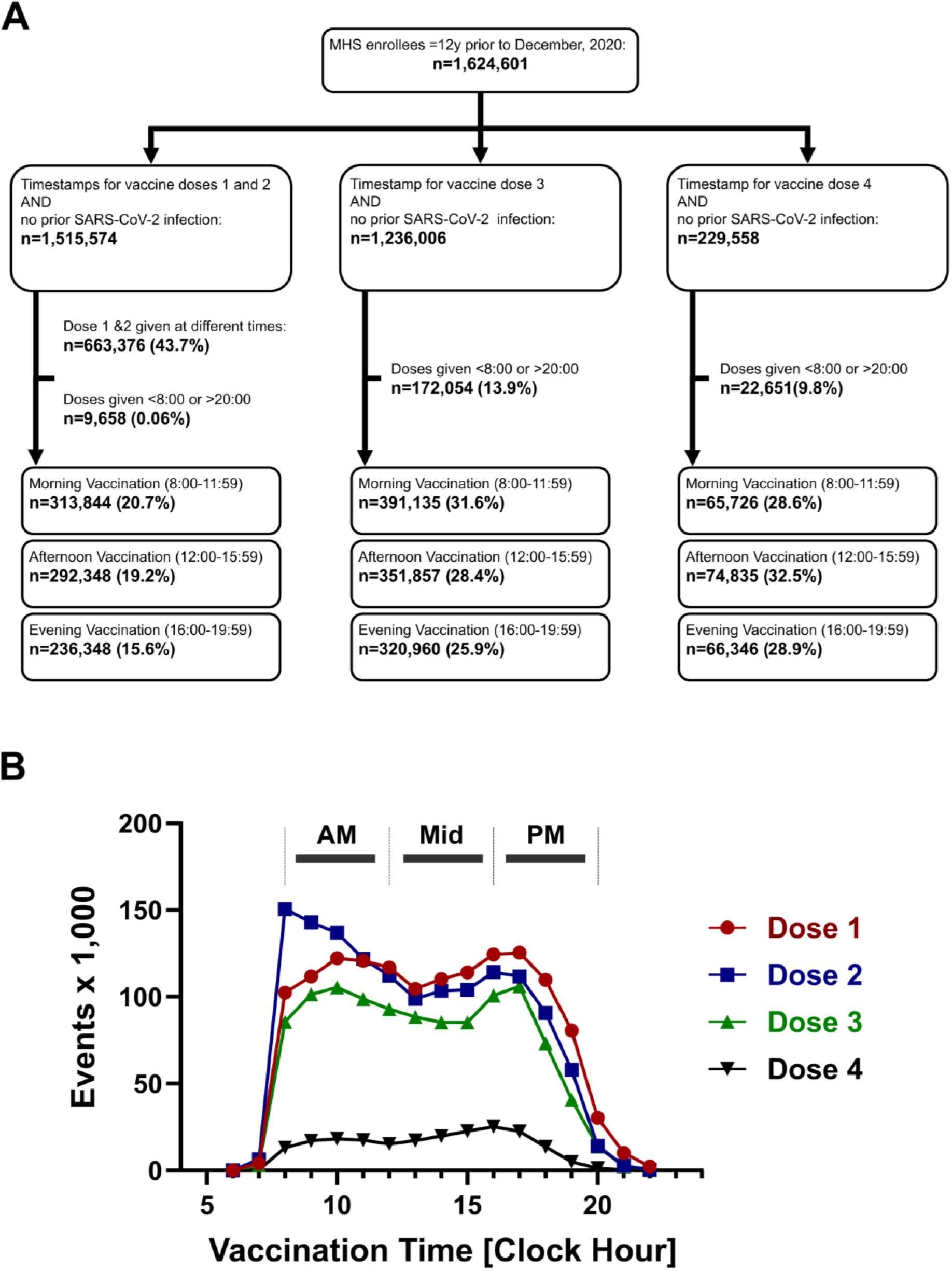
SARS-CoV2 vaccine timing across the day. **Panel A**shows the patient inclusion flowchart. **Panel B** shows the distribution of COVID-19 vaccine administration times, binned by hour of the day. Red circle, dose 1. Blue square, dose 2. Green triangle, dose 3 (the first booster dose). Black inverted triangle, dose 4 (second booster dose). Time intervals used to compare the effects of morning (AM), afternoon (Mid), and evening (PM) vaccine dosing on effectiveness are denoted by horizontal bars.

### Study population (Figure 1A)

We extracted data from all MHS members aged 12 years and older who received at least one dose of SARS-CoV-2 vaccine, joined MHS prior to February 2020 and therefore had a complete medical history on file. For analysis of each vaccine dose, we excluded patients who had a documented SARS-CoV-2 positive test prior to the date of vaccination. We further excluded patients with missing timestamps (11.4% of individuals for dose 1, 9.6% for dose 2, 12.3% for dose 3, and 9.1% for dose 4). 81.1% of MHS members identified as Jewish, 6% identified as orthodox Jews, 5.8% as Arabs, and 6.1% as former Soviet Union residents.

### Data sources and organization

We analyzed de-identified patient-level data extracted from MHS electronic records. Continuous variables included age at immunization, time and date of vaccine administration, and body mass index (BMI). Dichotomous variables included sex and comorbidities linked to COVID-19 severity^32,33^. For variable definitions see **eTables 1 and 2** in the **Supplement**.

**Table 1:** patient demographics based on time of COVID-19 vaccinations.^a^.

|  | **Doses 1&2<sup>b</sup>** |  |  | **Dose 3** |  |  | **Dose 4** |  |  |
| --- | --- | --- | --- | --- | --- | --- | --- | --- | --- |
| Variable | Evening,<br>N =<br>236,348 | Mid,<br>N =<br>292,278 | Morning,<br>N =<br>313,844 | Evening,<br>N =<br>320,960 | Mid,<br>N =<br>351,857 | Morning,<br>N =<br>391,135 | Evening,<br>N =<br>66,346 | Mid,<br>N =<br>74,835 | Morning,<br>N =<br>65,726 |
| **Age, Median (SD)** | 40 (19) | 42 (20) | 47 (20) | 44 (19) | 47 (19) | 46 (18) | 66 (12) | 69 (11) | 69 (11) |
| **Sex, n (%)** |  |  |  |  |  |  |  |  |  |
| F | 121,439 (51) | 155,133 (53) | 162,701 (52) | 165,086 (51) | 183,797 (52) | 205,816 (53) | 31,213 (47) | 37,990 (51) | 32,458 (49) |
| M | 114,909 (49) | 137,145 (47) | 151,143 (48) | 155,874 (49) | 168,060 (48) | 185,319 (47) | 35,133 (53) | 36,845 (49) | 33,268 (51) |
| **Comorbidities, n (%)** |  |  |  |  |  |  |  |  |  |
| 0 | 132,716 (56) | 162,156 (55) | 169,925 (54) | 175,356 (55) | 189,793 (54) | 211,664 (54) | 26,010 (39) | 29,877 (40) | 26,338 (40) |
| 1-4 | 100,865 (43) | 125,236 (43) | 137,278 (44) | 140,977 (44) | 155,506 (44) | 172,352 (44) | 36,989 (56) | 40,388 (54) | 35,402 (54) |
| 4+ | 2,767 (1.2) | 4,886 (1.7) | 6,641 (2.1) | 4,627 (1.4) | 6,558 (1.9) | 7,119 (1.8) | 3,347 (5.0) | 4,570 (6.1) | 3,986 (6.1) |
| **Diabetic, n (%)** | 17,576 (7.4) | 26,354 (9.0) | 33,157 (11) | 28,209 (8.8) | 34,693 (9.9) | 37,803 (9.7) | 16,252 (24) | 19,474 (26) | 17,313 (26) |
| **Blood Pressure, n (%)** | 38,122 (16) | 55,715 (19) | 72,071 (23) | 61,127 (19) | 75,120 (21) | 82,732 (21) | 33,129 (50) | 41,472 (55) | 35,964 (55) |
| **Neurological Disease, n (%)** | 20,415 (8.6) | 31,125 (11) | 36,610 (12) | 31,197 (9.7) | 39,046 (11) | 42,172 (11) | 13,422 (20) | 17,983 (24) | 15,757 (24) |
| **Chronic Kidney Disease, n (%)** | 8,828 (3.7) | 15,127 (5.2) | 20,011 (6.4) | 14,491 (4.5) | 20,074 (5.7) | 21,967 (5.6) | 9,736 (15) | 14,203 (19) | 11,994 (18) |
| **Immuno-suppression, n (%)** | 4,223 (1.8) | 6,191 (2.1) | 7,758 (2.5) | 6,640 (2.1) | 8,587 (2.4) | 9,169 (2.3) | 3,208 (4.8) | 3,855 (5.2) | 3,467 (5.3) |
| **Dialysis, n (%)** | 90 (<0.1) | 176 (<0.1) | 236 (<0.1) | 177 (<0.1) | 249 (<0.1) | 283 (<0.1) | 118 (0.2) | 144 (0.2) | 118 (0.2) |
| **Asthma, n (%)** | 43,420 (18) | 52,849 (18) | 52,543 (17) | 55,883 (17) | 61,236 (17) | 65,317 (17) | 10,146 (15) | 11,400 (15) | 10,199 (16) |
| **Cancer last 5yrs, n (%)** | 10,189 (4.3) | 16,340 (5.6) | 20,885 (6.7) | 16,497 (5.1) | 22,127 (6.3) | 23,681 (6.1) | 9,395 (14) | 12,403 (17) | 10,877 (17) |
| **Obesity, n (%)** | 43,167 (18) | 56,137 (19) | 64,610 (21) | 63,978 (20) | 70,721 (20) | 80,405 (21) | 19,427 (29) | 20,592 (28) | 18,279 (28) |
| **Heart Disease, n (%)** | 16,524 (7.0) | 25,474 (8.7) | 32,705 (10) | 25,691 (8.0) | 33,114 (9.4) | 36,730 (9.4) | 15,461 (23) | 19,800 (26) | 17,348 (26) |
| **Days between vaccinations 2-3, Median (SD)** | 197 (28) | 197(28) | 196 (27) |  |  |  |  |  |  |

<sup>a</sup>With the exception of sex, all differences between groups achieved statistical significance ( $p < 0.001$ , Wilcoxon rank sum test/Pearson's Chi-square test). This is a function of the large sample sizes in our cohort.
<sup>b</sup>Both vaccine doses administered within the indicated timeframe.

### Study outcomes

The primary outcome was COVID-19 infection as defined by positive SARS-CoV-2 polymerase-chain-reaction (PCR) or antigen test performed at any official site. Infections in the first 14 days following vaccination were excluded from the analysis, since patients during this interval are not considered to have a complete immunologic response to vaccination. The secondary outcome was emergency department (ED) visits associated with COVID-19 infection, defined as ED encounters in a period 7 days prior to 14 days after documented COVID-19 infection. We calculated number need to treat (NNT) as described^34^:

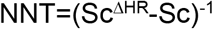

Where Sc is event free survival in the reference group and ΔHR is hazard ratio at the most minus the least preferable vaccination time. We estimated Sc as 0.75 at study end based on the survival curve generated for univariate analysis (**Figure 2**), and ΔHR as the peak-to-trough difference in HR for breakthrough infection across the day (**Figures 3 and 4**). We estimated the change in vaccine effectiveness based on moving vaccinations from the best to worst time-of-day as 1-HR as described^35^.

**Figure 2.**
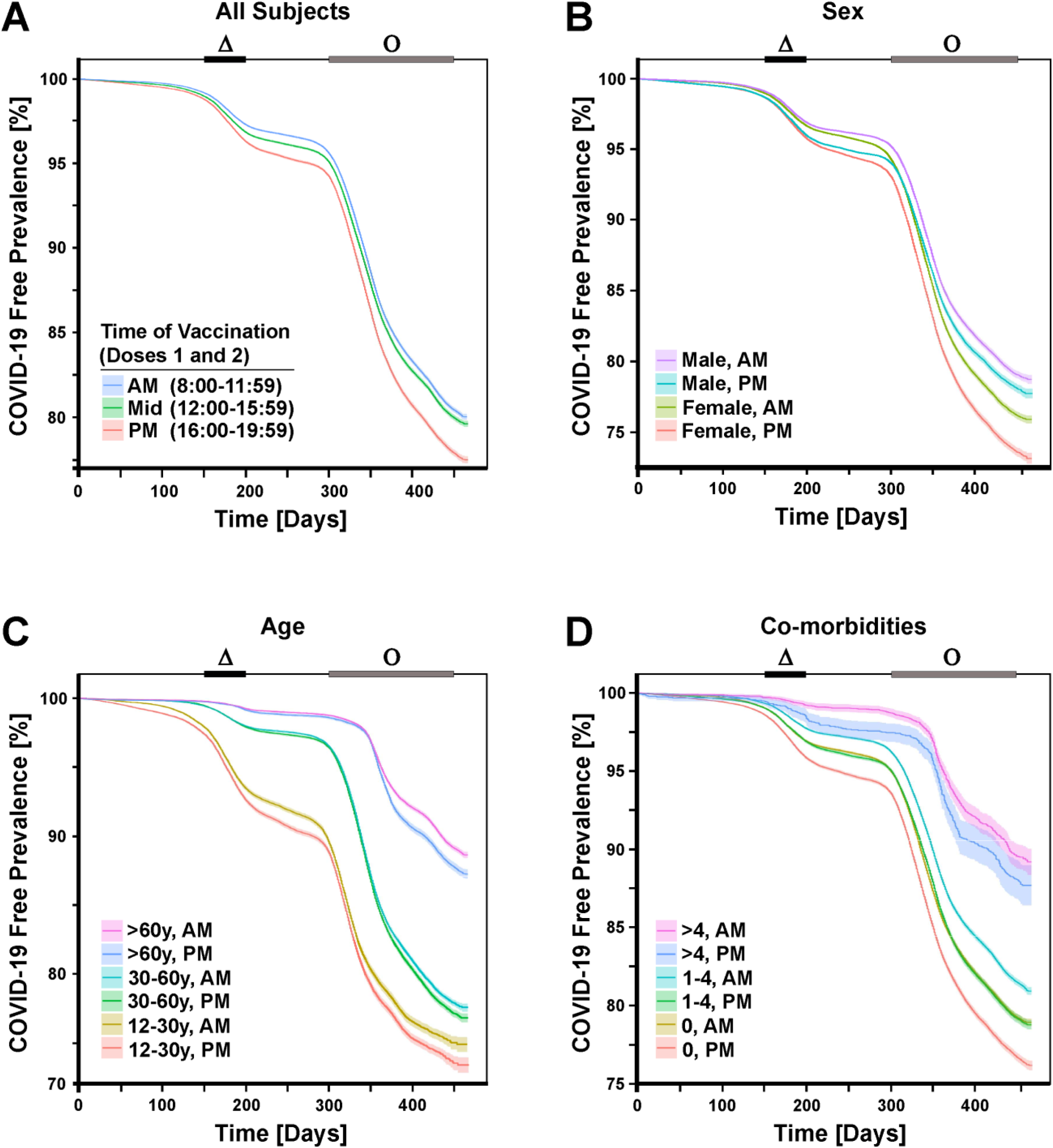
Morning COVID-19 vaccination is associated with fewer breakthrough infections. **Panel A** shows infection-free survival in patients receiving the first two COVID-19 vaccine inoculations between 8:00-11:59 hours (morning or AM), 12:00-15:59 hours (afternoon or Mid), and 16:00-19:59 hours (evening or PM). **Panels B-D** show COVID-19-free survival in patients stratified by sex (**B**), age (**C**), and number of medical co-morbidities (**D**). Shading around the lines represent 95% confidence intervals (CI). Waves of COVID-19 infection caused by the delta (Δ) and omicron (O) SARS-CoV-2 variants based on Israeli Ministry of Health (MOH) data are indicated by black and grey horizontal bars, respectively. For clarity, **panels B-D** display only the AM and PM groups.

**Figure 3.**
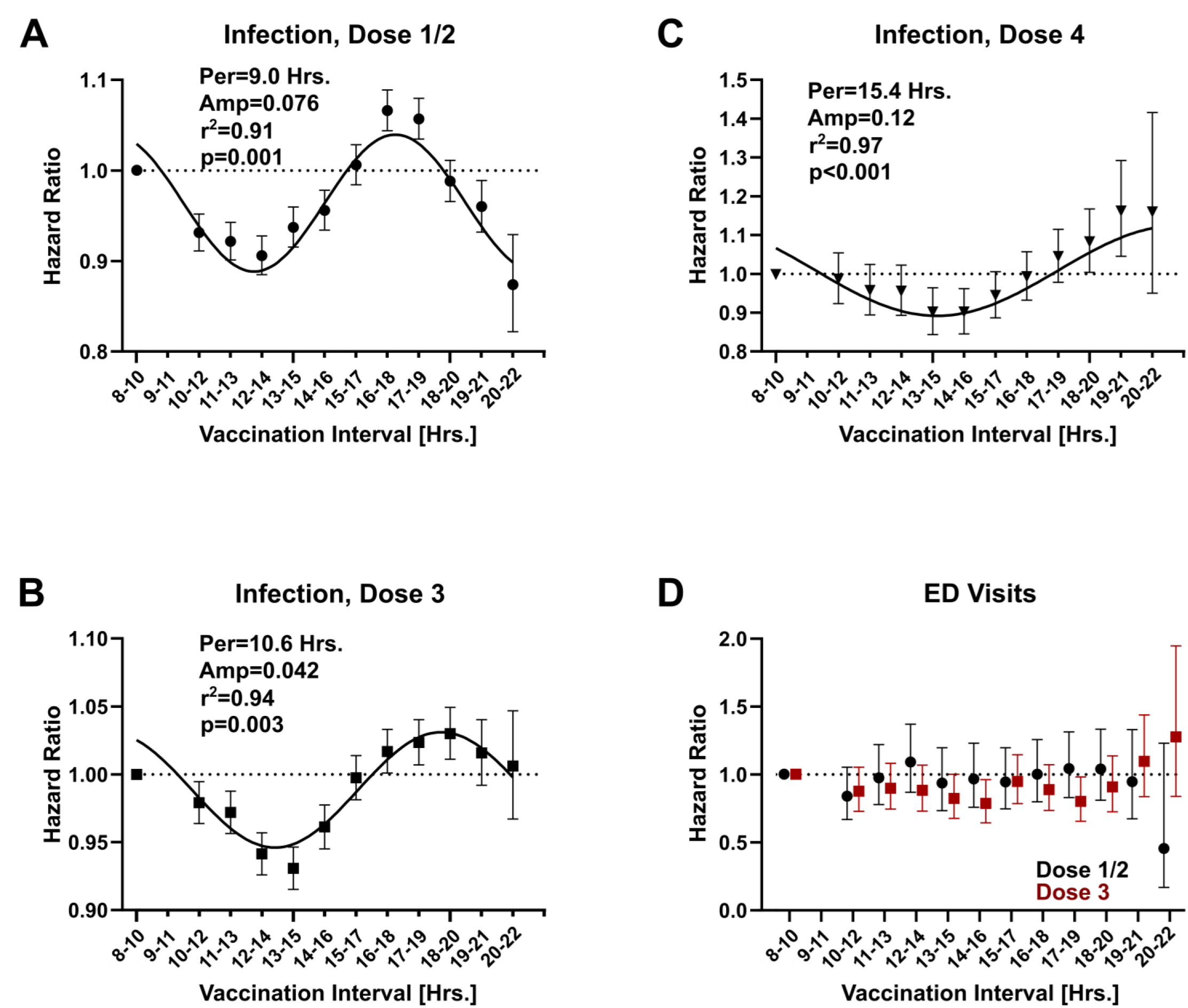
Biological rhythms in SARS-CoV-2 vaccine effectiveness. Data points represent adjusted hazard ratios (HR) ± 95% CI relative to dosing between 8:00-9:59 AM, which serves a non-overlapping index bin for this analysis. Best-fit sinusoidal trend lines (black lines), amplitudes (Amp), period duration (Per), goodness of fit (r^2^), and METACYCLE generated p-values for periodicity are depicted within each graph where appropriate. **Panels A-C** show HRs for breakthrough infections based on the timing of vaccine doses 1 and 2 (**A**), dose 3 (**B**), and dose 4 (**C**). **Panel D** shows HRs for COVID-19-associated ED visits, defined as occurring within -7 to +14 days of a COVID-19 positive test (see Methods). For patient demographic breakdown, sample sizes, and a tabular presentation of these data see **eTables 10-12** in then **Supplement**. For the complete METACYCLE output see **eTable 13** in the **Supplement**.

**Figure 4.**
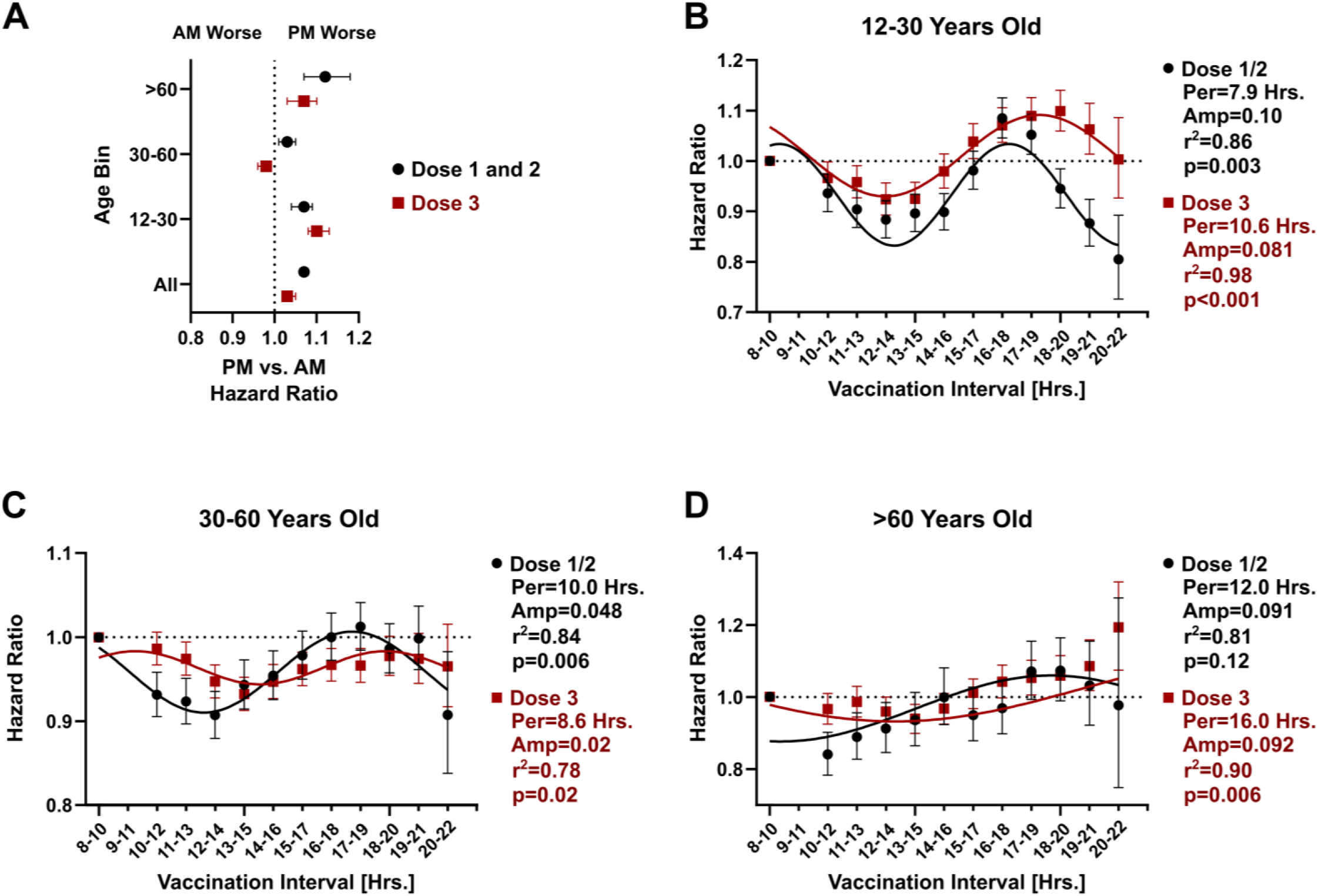
SARS-CoV-2 vaccine diurnal rhythms depend on patient age. **Panel A** shows HRs ± 95% CI for breakthrough COVID-19 infection based on time of vaccination (AM versus PM) and age range. Values to the right of the dotted line favor morning vaccination (8:00-11:59), and values to the left favor evening vaccination (16:00-19:59). Black, timing of doses 1 and 2 are considered. Red, timing of dose 3 is considered. **Panels B-D** show comparative vaccine effectiveness around the clock relative to dosing between 8:00-9:59 (HR ± 95% CI). Best-fit sinusoidal trend lines (black lines), period duration (Per), amplitudes (Amp), goodness of fit (r^2^), and METACYCLE generated p-values for periodicity are depicted within each graph. Black, timing of doses 1 and 2 are considered. Red, timing of dose 3 is considered. (**B**) Ages 12-30 years old. (**C**) Ages 30-60 years old. (**D**) Over 60 years old. For sample sizes and tabular presentation of these data see **eTables 14-17** in the **Supplement**. For the complete METACYCLE output see **eTable 13** in the **Supplement**.

### Statistical analysis

An initial descriptive analysis included calculations of single variable distribution, central tendency, and dispersion. We stratified vaccine timing in 4-hour bins: 8:00-11:59 (morning), 12:00-15:59 (afternoon), and 16:00-19:59 (evening) comparison groups (**Figure 1B**). Day 0 in our analysis was individually defined for each patient as 7 days after the receipt of vaccine dose 2 as described^36^ Given that the first 2 doses of BNT162b2 constitute a single intervention we combined these doses into one analytic model. Patients who received both doses 1 and 2 in the same time bin were included in this analysis, while patients vaccinated in different time bins were excluded (**Figure 1A**). For analysis of the booster doses (doses 3 and 4), only the time of booster administration was considered as boosters are regarded as separate medical interventions to augment waning COVID-19 immunity.

Since one independent variable (booster status) differed over time, univariate and multivariate survival analyses were performed with time-dependent covariates^37,38^. Kaplan–Meier analysis with a log-rank test was used for the univariate analysis. A Cox proportional-hazards regression model with time-dependent covariates was used to estimate the association between vaccine administration time-of-day and study endpoints. This statistical approach was previously applied to Israeli HMO data for analyzing associations between COVID-19 immunization and clinical endpoints over extended periods^35^. Our choice of variables to include in the Cox model was based on previously published associations between age, comorbidities and COVID-19 endpoints of infection or disease severity (see **eAppendix3** in the **Supplement** for a discussion of Cox model variable selection)^32,33,35^. To simplify the modeling of breakthrough events after the initial immunization series (doses 1 and 2), we excluded subjects who received the second booster (dose 4). For modeling events after doses 3 and 4 we included all subjects. We treated the first COVID-19 infection as a terminal event. As such, recurrent infections did not factor into our analysis. For orthogonal validation of conclusions from our Cox model and a discussion of proportional hazard assumptions and see “Model Sensitivity Analysis” in **eAppendix4 and eTables 3-5** in the **Supplement**.

To examine how the choice of time bins affected conclusions from our Cox regression model, we generated HRs comparing the 8:00-9:59 time bin to successive 2-hour intervals across the day, incrementing by one hour with each iteration. Note, to avoid overlap with the index bin, HRs were generated from 10:00 on in this analysis. Our hypothesis was that a diurnal rhythm in vaccine effectiveness should produce a sinusoidal trend in HRs as a function of vaccination time. To statistically evaluate the trend in HRs across the day for periodicity we used METACYCLE, an algorithm commonly used for circadian rhythm detection in gene expression data^39^. The COSOPT algorithm^40^ was used to generate best-fit sinusoidal curves for data presentation. We observed very high goodness of fit with this approach (r^2^ range 0.91-0.97, **Figure 3**), supporting the use of a sinusoidal model. To avoid forced fitting these data, we programmed COSOPT and METACYCLE to select the best fitting periodic function across a range of cycle lengths (4-24 hours) as described^40^.

We performed all statistical analyses using R version 3.5.0. Code for all analyses is available upon request. A p-value of less than 0.05 was used to indicate significance in all analyses.

## RESULTS

To examine how the timing of COVID-19 immunization relates to clinical effectiveness we analyzed a large cohort of patients enrolled in Maccabi Healthcare Services (MHS), a major Israeli HMO. A strength of MHS data is that prior studies have used it and other similar sources to measure the real-world effectiveness of COVID-19 vaccines^35,36,41,42^. However, these studies did not consider the time-of-immunization as a factor.

Out of approximately 2.6 million MHS participants, 1,515,574 had timestamps recorded for at least 1 immunization (**Figure 1A**). Our study population almost exclusively received the Pfizer-produced BNT162b2 vaccine (99.2%, n=1,503,599), with a minority receiving the Moderna mRNA-1273 product (0.74%, n=11,220). Most patients received immunizations within a 12-hour timespan stretching from 8:00 to 20:00 (**Figure 1B**). Based on this distribution, we began by comparing COVID-19 breakthrough infections in patients receiving their first two immunizations exclusively in the morning (8:00-11:59, n=313,844), afternoon (12:00-13:59, n=292,278), or evening (16:00-19:59, n=236,348). As a group, patients vaccinated in the morning were older and had more comorbidities than at other times (**Table 1**).

Patients immunized against COVID-19 in the morning and afternoon had less frequent breakthrough infections than those vaccinated in the evening, with the groups diverging prior to the wave of infections caused by the Delta variant (**Figure 2A**). The difference remained significant after stratification by sex, age, and total number of co-morbidities (**Figure 2B-D**).

Counterintuitively, factors like older age that predict worse COVID-19 clinical outcomes were associated with fewer breakthrough infections, a finding also noted elsewhere^35,41^. We suggest this reflects greater adherence to COVID-19 precautions in older or more vulnerable individuals (for example, mask use and social distancing), leading to less viral exposure. Even so, morning vaccination remained superior to evening vaccination in these patients (**Figures 2C, D**).

We considered whether patients receiving vaccinations at different times of day might have different baseline infectious risks due to unmeasured variables in our data, for example occupation or household size. To this end, we scrutinized COVID-19 infections in the first 14 days after the first immunization, a timeframe prior to the onset of full vaccine protection. During this period there were 7.0, 8.1, and 3.6 COVID-19 infections per 100,000 patients in the morning, afternoon, and evening groups respectively (p=0.008 morning vs. evening and p=0.245 morning vs. afternoon, permutation test). This pattern is the opposite of the effectiveness signal seen after vaccination takes effect, favoring morning or afternoon dosing (**Figure 2A**). Since patients vaccinated in the morning are older and have more comorbidities (**Table 1**), adjustment for these demographic variables would further widen the difference between morning and evening groups. Thus, baseline infection risk does not explain why morning and afternoon vaccinations were associated with fewer breakthrough infections post-immunization.

We also considered if our comparison groups might differ in their readiness to undergo COVID-19 testing, thus confounding results. However, the morning and evening vaccination groups had equivalent numbers of COVID-19 tests (2.28 vs. 2.29 tests per capita, respectively) and a similar distribution of testing times around the clock (**eFigure 2A** in the **Supplement**). We did observe reduced positivity rates in COVID-19 tests obtained in the early morning like one recent report^43^, but this pattern was independent of vaccination timing (**eFigure 2B** in the **Supplement**). Overall, univariate analysis suggested a clinical advantage to dosing COVID-19 vaccines in the morning or afternoon in terms of fewer breakthrough infections. These results are unlikely to reflect differences in the baseline velocity of COVID-19 infection, diagnostic testing patterns, or diurnal variations in COVID-19 detection.

Using Cox multivariate regression to adjust for age, sex, comorbidities, and the protection afforded by COVID-19 boosters, we mapped the relationship between vaccination time and breakthrough infections (**Figure 3**). Across the 14-hour span when most COVID-19 vaccinations were given, the hazard ratio (HR) for breakthrough infections was lowest between late morning to early afternoon and highest risk with evening vaccination times (**Figure 3A-C**). Stratifying subjects into morning, mid-day, and evening vaccination groups produced similar conclusions (**eTables 6-8** in the Supplement), as did the use of two alternative statistical approaches: logistic regression and bootstrap analysis (**eTables 4 and 5** in the **Supplement**). The relationship between vaccination time and HR for breakthrough infection was sinusoidal with period durations ranging from 9.0-15.4 hours, depending on the vaccine dose (**Figure 3A-C**). We estimated a maximum peak-to-trough change in the HR for breakthrough infection as 0.12 for doses 1 and 2, 0.086 for dose 3, and 0.25 for dose 4 (**Figure 3A-C**). In our cohort, this translates to a number needed to treat (NNT) range of 18.7-54.5 by study end if patients were moved from the least favorable to most favorable COVID-19 vaccination times. It also corresponds to a relative change of vaccine effectiveness of 8.6-25%. In contrast to breakthrough infections, vaccine timing was not associated with rates of COVID-19 associated ED visits, although these encounters correlated with age and comorbidities as previously described^32,33^ (**Figure 3D and eTable 9** in the **Supplement**). Thus, COVID-19 vaccination timing had a significant association with effectiveness as defined by breakthrough infection, but this did not translate directly into ED visits. The sinusoidal trend suggests a biological rhythm in COVID-19 vaccine effectiveness based on time of administration.

Since age affects breakthrough COVID-19 infection rates in our data (**Figure 2C and eTables 6-8** in the **Supplement**), we examined how this variable interacted with vaccine effectiveness (**Figure 4**). We found that the benefits of morning vaccination were concentrated in younger (<30 years old) and older (>60 years old) individuals (**Figure 4A**). For both the initial vaccine series and the first booster (dose 3), increasing age correlated with longer period durations and a shift in the peak HR to later hours, perhaps reflecting changes to the circadian system and immune experience with aging (**Figure 4B-D**)^44,45^. In the oldest age stratification (**Figure 4D**), the rhythms resembled that of the second booster (dose 4, **Figure 3C**), an intervention restricted to older patients during the study period (**Table 1**). Taken together, these data suggest a strategy for leveraging diurnal rhythms to optimize COVID-19 vaccine effectiveness in a population, where younger patients and the elderly would be prioritized for immunizations in the late morning to early afternoon.

## DISCUSSION

Our data indicate a significant association between the time of COVID-19 vaccination and its clinical effectiveness in terms of breakthrough infection. While the impact of vaccination timing is additive on top of an effective vaccine, it is an easily modifiable factor that when extended over millions of immunizations can amount to a large aggregate benefit. The relationship between vaccination timing and infection risk is sinusoidal, suggesting a biological rhythm in vaccine effectiveness consistent with circadian regulation of underlying immune processes. While breakthrough COVID-19 infections varied with vaccination timing, ED presentations proximate to COVID-19 detection did not. This finding may indicate that while COVID-19 vaccine immunogenicity is circadian regulated, patient factors like age and co-morbidities dominate the clinical severity of disease. Alternatively, our statistical power for detecting associations with ED presentations was lower as they were infrequent in our cohort (one ED visit to 60.8 positive COVID tests). Regardless, minimizing COVID-19 infections of any severity is desirable. Asymptomatic patients can still infect others and even mild symptoms can complicate the management of chronic conditions. Our observation that biological rhythms map to younger patients and the elderly is important because it suggests that prioritizing these patients for mid-day immunization could improve the effectiveness of mass vaccinations against COVID-19. We suggest this idea is practical and warrants prospective study.

Human populations have less consistent behavioral patterns than other organisms, making it challenging to observe how circadian rhythms affect medical interventions in real-world settings. This is because individuals lead diverse lifestyles, are routinely exposed to light at night, and engage in activities like night shift work that alter the phase of the circadian clock^19,46^. Our ability to detect rhythms in COVID-19 vaccine effectiveness in this study is due to the unique circumstances of the COVID-19 pandemic and the Israeli response to the pandemic. This includes a population-level sample of subjects that is two orders of magnitude larger than prior cohorts, a concerted intervention largely consisting of a single vaccine product (BNT162b2), a population highly motivated to report COVID-19 infection, substantial clinical follow-up (>1 year), and finally a high level of data integration within the Israeli healthcare system. Our data do not directly address whether rhythms in BNT162b2 effectiveness extend to other COVID-19 vaccine products or to vaccinations against other pathogens. However, it is worth noting that the plurality of studies that do report diurnal rhythms in vaccine immunogenicity found morning to afternoon dosing to be optimal^23,27,28^. Therefore, our data may have implications for diseases beyond COVID-19.

Our analysis has limitations. As with any observational study, patients were not randomly assigned to specific vaccination times, and demographic differences between groups can bias results. We attempted to compensate for this by adjusting for variables known to affect COVID-19 infection rates and complications^32,33,35^, assessing the role of unmeasured variables, and using independent statistical models. Nevertheless, there may be sources of bias missed by our methods. There were also a limited number of ED events in the study population to support conclusions related to this specific outcome. Our dataset lacks comprehensive viral genotyping and so we cannot determine whether time-of-vaccination influences breakthrough infections to different extents across variants. However, given rhythms were evident with vaccine boosters given while different viral variants were dominant our findings are likely to be broadly applicable. Because vaccinations were not provided around the clock in our population, we cannot be certain whether the observed rhythms in vaccine effectiveness are diurnal (i.e., 24-hour cycles) or ultradian in nature (<24 hours cycles) as suggested by periodicity analysis (**Figures 3 and 4; eTable 13** in the **Supplement**). Regardless, both diurnal and ultradian rhythms are compatible with upstream circadian clock influence^18,47^. Our results reflect the cumulative behavior of our cohort, and individuals with acutely disrupted circadian rhythms due to shift work, jet lag or sleep deprivation might react differently to immunization timing. Research suggests these patients are more vulnerable to COVID-19 infection and future research should focus on vaccination strategies specific to this subgroup^48^. In summary, circadian rhythms represent a fundamental property of living things that regulate immune function at a basic biological level. This study provides the first estimate of how far this core process extends into an important clinical realization of immune function: vaccine effectiveness. Our data suggest a way of leveraging biological rhythms to optimize immunizations against SARS-CoV-2 variants using reformulated vaccines and potentially vaccines against other pathogens.

## Data Availability

R code for all analyses is available upon request. Pursuant to Maccabi Healthcare Services data sharing policy, individual patient data will not be shared. We provide a full list of outcome variables and clinical covariates for this study in eTables 1 ans 2 of the Supplemement.

## DATA SHARING STATEMENT

R code for all analyses is available upon request. Pursuant to MHS data sharing policy, patient data will not be shared.

## ACKNOWLEDGEMENTS

We thank Steven Brody, Erik Herzog, Patrick Lyons, Edward Puro, and Robyn Haspel for their input.

