## Supplemental methods, figures, and tables. for "BIOLOGICAL RHYTHMS IN COVID-19 VACCINE EFFECTIVENESS"

**eAppendix 1**

**Study Oversight.**

Dr. Hazan and Dr. Haspel conceived the study. All the authors contributed to the study design. Data were extracted by Hillel Alapi. Dr. Hazan, Dr. Duek, and Gabriel Chodick oversaw the data gathering. All authors contributed to the data analysis. Dr. Hazan and Dr. Haspel wrote the initial manuscript. All authors vouch for the data and analysis. All authors participated in the editing and critical review of the manuscript and decided to submit it for publication. The study was approved by the MHS Ethical Committee.

**eAppendix 2**

**Background on COVID-19 vaccinations in Israel.**

In December 2020, Israel launched a national immunization campaign against COVID-19 consisting almost exclusively of the Pfizer BNT162b2 vaccine series delivered in two doses 3-4 weeks apart (**Figure S1**). In July and September 2021, the Israeli Ministry of Health initiated third (first booster) and fourth (second booster) dose campaigns, respectively. Vaccines and COVID-19 testing were provided free of charge to all citizens. While at-home antigen tests were available, the Israeli Ministry of Health required a documented positive test prior to quarantine and a negative test after quarantine before issuing a certificate of recovery to patients (https://corona.health.gov.il/en/confirmed-cases-and-patients/cases-recovered/). Thus, censoring of COVID-19 infection is unlikely in our cohort as there was strong incentive to obtain a laboratory diagnosis even if initial testing occurred at home.

Among 1,624,601 MHS participants over 12 years of age 1,594,180 (98.13%) received the Pfizer BNT162b2 vaccine, 29,253 (1.8%) received the Moderna mRNA-1273 vaccine, 437 (0.03%) received the Astra Zeneca ChAdOx1-S product, 400 (0.02%) received Sputnik V, 232 (0.01%) received the Jansen Jcovden vaccine, 46 (0.003%) received the Sinovac CoronaVac product, and for 53 patients (0.003%) the vaccine identify was unspecified.

**eAppendix 3**

**Selection of variables for Cox multivariate regression.**

We conducted multivariate cox regression model based on clinical relevance and univariate comparison. As a starting point, we included variables who known to be associated with vaccine effectiveness based on previous publications[1-3]. Then, we interrogated violation of Cox assumptions for the variables of interest by computing a Schoenfeld global test (see "Cox model sensitivity analysis" in the Supplementary Appendix below). Variables that were associated with outcomes measured (breakthrough COVID-19 infection or COVID-19-related emergency room visit) with a $p<0.1$ in the univariate comparison were included in the multivariate model. For the multivariate analysis we used goodness of fit parameters (AIC – Akaike Information Criterion = -2 Log likelihood+ 2p) with a backward elimination approach.

**eAppendix 4**

**Cox model sensitivity analysis**.

Typically, a Schoenfeld's global test is used to validate the assumption of Cox proportional hazards. However, the large sample sizes of our study present a challenge to this approach since this leads to over-powering[4]. We found evidence of this in our data by applying the Schoenfeld's global test on stratified data with smaller sample sizes (**Table S3**), demonstrating reduction in test significance compared to the complete dataset. Therefore we used two alternate approaches to orthogonally validate results from our Cox model as recommended[4]: multivariate logistic regression (**Table S4**) and bootstrap multivariate analysis (**Table S5**). For logistic regression, we applied the same multivariate Cox regression model, with the exception that follow-up time was not included. The reason for this modification was our goal with logistic regression was to validate the trend rather than the effect size estimated by the Cox model, and that follow-up times were not materially different between vaccination groups (**Table 1**). For bootstrap analysis, we sampled the entire cohort with 2,000 iterations with replacement calculating hazard ratios and confidence intervals for the same multivariate regression model. Results from both approaches showed similar trends as our multiple Cox regression model, supporting this approach for analyzing the data. As additional support, a plot of scaled Schoenfeld residuals for the variable time-of-vaccination demonstrated stable proportional hazard ratios across the study period (**Fig. S3**)[5].

eFigure 1

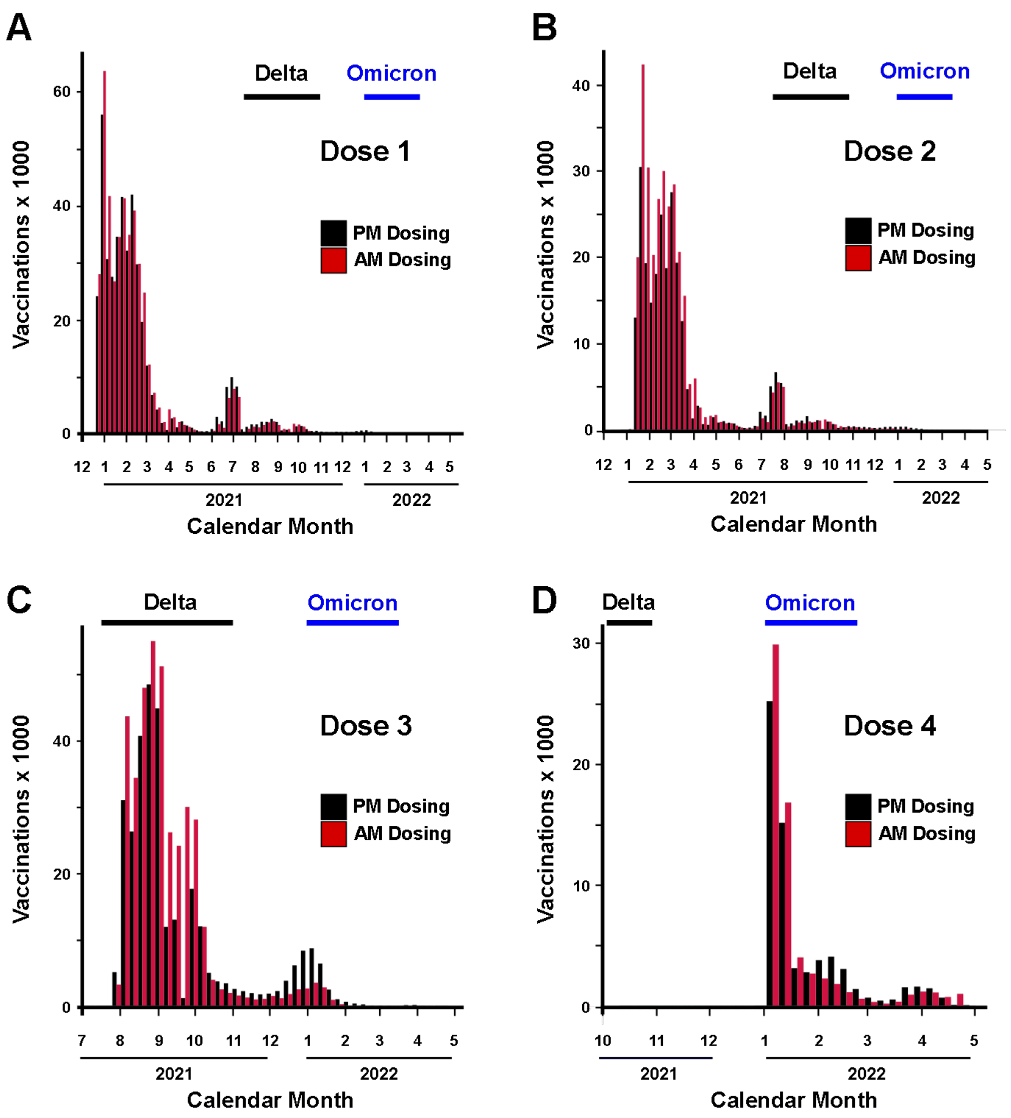

**eFigure 1. COVID-19 vaccination schedule in the MHS cohort.** Bars represent the number of vaccines given between 8:00-11:59 (AM dosing, red bars) and 16:00-19:59 (PM dosing, black bars). Calendar months are depicted numerically with 1 representing January and 12 representing December. Spikes in infection rates within the Israeli population caused by the delta and omicron SARS-CoV-2 variants as reported by the Israeli Ministry of Health (MOH) are indicated by black and blue horizontal bars, respectively. **Panels A-D** show vaccine doses over time for the first (**A**), second (**B**), first booster (third dose, **C**), and second booster doses (fourth dose, **D**).

### eFigure 2

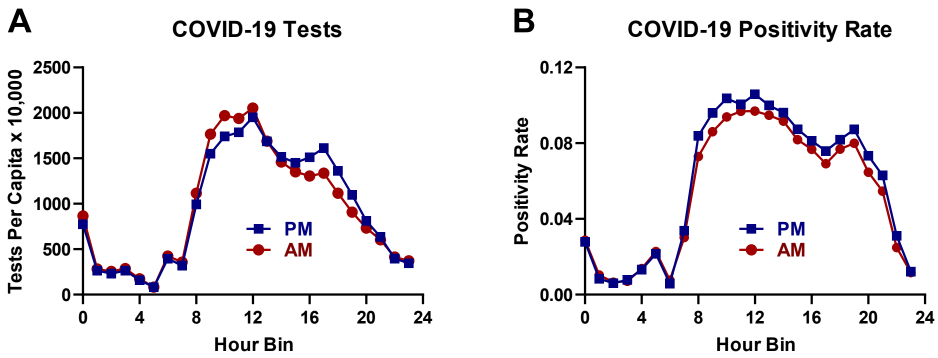

**eFigure 2. Distribution of COVID-19 positivity across the day as a function of COVID-19 vaccine timing**. **Panel A and B** show the distribution of total COVID tests (**A**) and positivity rates (**B**) across the day for patients receiving the initial COVID-19 vaccine series (doses 1 and 2). AM, patients immunized between 8:00-11:59 (red symbols). PM, patients immunized between 16:00-19:59 (blue symbols). For sample sizes and tabular presentation of these data see **eTable 18**.

**eFigure 3**

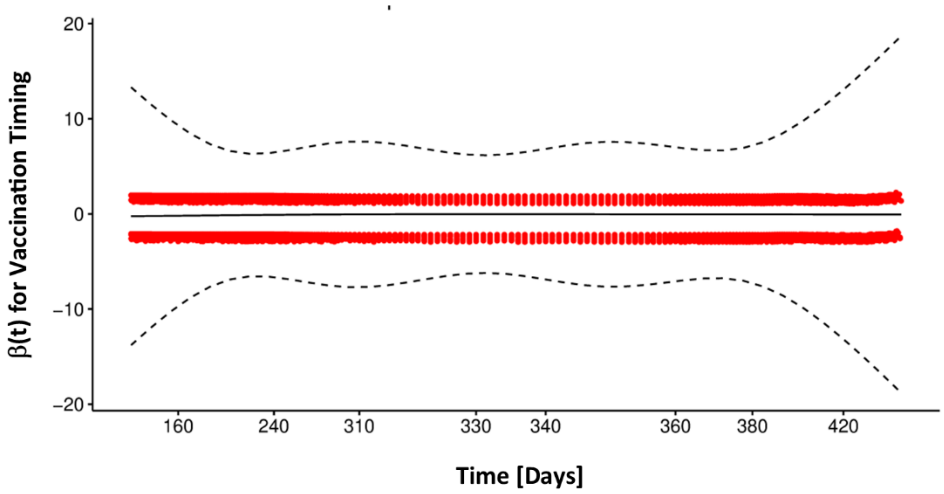

**eFigure 3. Plot of scaled Schoenfeld residuals for vaccination timing over the study period.** Dashed lines, 95% CI. For discussion of Cox proportional hazards assumption, see “Covid Model Sensitivity Analysis” in the Supplemental Appendix.

**eTable 1: Outcome variable definitions.** Note, we report all outcome variables extracted for this project regardless of whether they entered into the analysis reported by this manuscript.

| Variable | Values | Definitions | Timing |
|---|---|---|---|
| Documented SARS-CoV-2 Infection | 0/1 | A PCR confirmed infection. | The date and time-of-day of a specimen collection that was found to be positive in a PCR test.<br><br>If the PCR test was done after the beginning of a hospitalization flagged as a COVID-19 hospitalization, the infection date was set to the beginning of the hospitalization. |
| Documented SARS-CoV-2 Infection | | A PCR: Cycle threshold number. | |
| Asymptomatic SARS-CoV-2 Infection | 0/1 | A PCR-confirmed infection with no report of symptoms during referral and during initial physician questioning. | The date set for theSARS-CoV-2 infection outcome. |
| COVID-19 (symptomatic SARS-CoV-2 Infection) | 0/1 | A PCR-confirmed infection with report of symptoms during the PCR referral / during the follow-up in the community setting / COVID-19 related hospitalization / COVID-19 related death.<br><br>Existing symptoms were considered when the physician or nurse checked the "symptomatic" option in the EMR, or when the following specific symptoms were recorded: fever or chills, cough, shortness of breath or difficulty breathing, sore throat, headache, weakness, congestion or runny nose, myalgia, nausea or vomiting, diarrhea, abdominal pain, loss of taste or smell,inability to eat or drink. | The date set for theSARS-CoV-2 infection outcome. |

| | | | |
|---|---|---|---|
| COVID-19 related emergency room visit | 0/1 | | Date and time-of-day |
| COVID-19 related hospitalization | 0/1 | A hospitalization that was reported to the Israeli Ministry of Health (MOH) as a hospitalization of a SARS-CoV-2 infected individual. | The start date of the hospitalization. |
| COVID-19 related severe state | 0/1 | As defined by the hospitalizing institution per the Israeli MOH guidelines, consistent with the NIH criteria for severe illness or critical illness: Individuals who have oxygen saturation ($SpO_2$) <94% on room air at sea level, a ratio ofarterial partial pressure of oxygen to fraction of inspired oxygen ($PaO_2/FiO_2$) <300 mm Hg, respiratory frequency >30 breaths/min, or lung infiltrates >50%.<br><br>Critical Illness: Individuals who have respiratory failure, septic shock, and/or multiple organ dysfunction. | The first date during the hospitalization in which the individual was flagged as being in a severe or critical state. |
| COVID-19 related death | 0/1 | A death of a SARS-CoV-2 infected individual reported to the Israeli MOH. | The reported date of death. |
| COVID-19 vaccine-related side effect | 0/1 | | |
| SARS-CoV-2-IgG serology post COVID-19 vaccine | 0/1 | Level of SARS-CoV-2-IgG | Date of the test and time-of-day |

**eTable 2: Clinical covariate definitions.**

| Variable | Values | Definitions | Timing |
|---|---|---|---|
| Age | | Age in complete years | Current |
| Sex | Male/Female | As defined in files | Current |
| COVID Vaccine | | Dose number, type | Date for each dose |
| COVID Vaccine | | | Time-of-day for each dose |
| Health-care worker | 0/1 | Is patient a health-care worker per File. | Current |
| Long-term care facility resident | 0/1 | Is patient a long-term care facility resident per file. | Current |
| Cancer | 0/1 | ICD9 Code 174*<br>ICD9 Code 175*<br>ICD9 Code 233.0<br>ICD9 Code V10.3<br>ICD9 Proc Code 85.4*<br>ICD9 Code 153*<br>ICD9 Code 154*<br>ICD9 Code V10.5*<br>ICD9 Code V10.6*<br>ICD9 Code 185<br>ICD9 Code V10.46<br>ICD9 Code 162*<br>ICD9 Code V10.1*<br>ICD9 Code 188*<br>ICD9 Code V10.51<br>ICD9 Code 183*<br>ICD9 Code V10.43<br>ICD9 Code 179<br>ICD9 Code 182*<br>ICD9 Code V10.42<br>ICD9 Code 157*<br>ICD9 Code 191*<br>ICD9 Code 192*<br>ICD9 Code V10.85<br>ICD9 Code 151*<br>ICD9 Code V10.04<br>ICD9 Code 172*<br>ICD9 Code V10.82<br>ICD9 Code 201*<br>ICD9 Code 200*<br>ICD9 Code 202.4*<br>ICD9 Code 204*<br>ICD9 Code 205*<br>ICD9 Code 206*<br>ICD9 Code 207.1*<br>ICD9 Code 208.1* | Last 5 years |

| | | | |
|---|---|---|---|
| | | ICD9 Code 189*<br>ICD9 Code V10.52<br>ICD9 Code 160*<br>ICD9 Code 161*<br>ICD9 Code 164.0<br>ICD9 Code 195.0<br>ICD9 Code V10.21<br>ICD9 Code V10.22<br>ICD9 Code 180*<br>ICD9 Code V10.41<br>ICD9 Code 140*<br>ICD9 Code 141*<br>ICD9 Code 142*<br>ICD9 Code 143*<br>ICD9 Code 144*<br>ICD9 Code 145*<br>ICD9 Code 150*<br>ICD9 Code V10.03<br>ICD9 Code 155*<br>ICD9 Code 156*<br>ICD9 Code V10.07<br>ICD9 Code 170*<br>ICD9 Code V10.81<br>ICD9 Code 193<br>ICD9 Code V10.87<br>ICD9 Code 171*<br>ICD9 Code 176*<br>ICD9 Code 184*<br>ICD9 Code 186*<br>ICD9 Code 187*<br>ICD9 Code V10.4*<br>ICD9 Code 203*<br>ICD9 Code 273.3<br>ICD9 Code 152*<br>ICD9 Code 158*<br>ICD9 Code 159*<br>ICD9 Code 163*<br>ICD9 Code 164*<br>ICD9 Code 165*<br>ICD9 Code 181<br>ICD9 Code 190*<br>ICD9 Code 192.8<br>ICD9 Code 196*<br>ICD9 Code 197*<br>ICD9 Code 198*<br>ICD9 Code 199* | |
| Chronic Kidney Disease | 0/1 | ICD Proc Code 39.95<br>ICD Proc Code 54.98<br>ICD9 Code 996.81 | Ever |

| | | | |
|---|---|---|---|
| | | ICD9 Code V42.0<br>ICD Proc Code 55.6*<br>ICD9 Code 403._1<br>ICD9 Code 404._2<br>ICD9 Code 404._3<br>ICD9 Code 585*<br>ICD9 Code 586<br>ICD9 Code 250.4*<br>ICD9 Code 274.1*<br>ICD9 Code 440.1<br>ICD9 Code 581*<br>ICD9 Code 582*<br>ICD9 Code 583*<br>ICD9 Code 587<br>ICD9 Code 588*<br>ICD9 Code 589* | |
| Chronic Obstructive Pulmonary Disease | 0/1 | ICD9 Code 491*<br>ICD9 Code 492*<br>ICD9 Code 496 | Ever |
| Heart Conditions | 0/1 | ICD9 Code 410*<br>ICD9 Code 411*<br>ICD9 Code 412<br>ICD9 Code 413*<br>ICD9 Code 414*<br>ICD9 Code 429.2, 429.7*<br>ICD9 Code V45.81, V45.82<br>ICD9 Proc Code 36.0*<br>ICD9 Proc Code 36.1*<br>ICD9 Code 428*<br>ICD9 Code 398.91<br>ICD9 Code 402._1<br>ICD9 Code 404._1,<br>ICD9 Code 404._3<br>ICD9 Code 416.9<br>ICD9 Code 514<br>ICD9 Code 425*<br>ICD9 Code 416* | Ever |
| Solid Organ Transplant Recipient | 0/1 | ICD9 Code 996.81<br>ICD9 Code V42.0<br>ICD Proc Code 55.6*<br>ICD9 Code V42.7<br>ICD Proc Code 50.5*<br>ICD9 Code V42.1<br>ICD9 Code V43.2<br>ICD Proc Code 37.5<br>ICD9 Code V42.83<br>ICD Proc Code52.8*<br>ICD9 Code V42.6 | Ever |

| | | | |
|---|---|---|---|
| | | ICD Proc Code33.5*<br>ICD Proc Code 33.6 | |
| Obesity | 0/1 | Body Mass Index (BMI) 30-40 | Latest measurement in last 5 years not taken during pregnancy |
| Severe Obesity | 0/1 | Body Mass Index (BMI) 40+ | Latest measurement in last 5 years not taken during Pregnancy |
| Pregnancy | 0/1 | | Current |
| Sickle Cell Disease | 0/1 | ICD9 Code 282.6* | Ever |
| Smoking | 0/1 | | Last recorded value |
| Type 2 Diabetes Mellitus | 0/1 | HbA1C > 6.5<br>ATC Codes A10[A,B]<br>ICD9 Code 250*<br>ICD9 Code 357.2<br>ICD9 Code 362.0*<br><br>**And not:**<br>ICD9 Code 250._1, 250._3 | For diagnosis codes, ever<br><br>For drugs, 4 or more dispensed in last 12 months |
| Asthma | 0/1 | ICD9 Code 493* | Ever |
| Cerebrovascular Disease | 0/1 | ICD9 Code 362.34<br>ICD9 Code 430<br>ICD9 Code 431<br>ICD9 Code 432*<br>ICD9 Code 433*<br>ICD9 Code 434*<br>ICD9 Code 435*<br>ICD9 Code 436*<br>ICD9 Code 438* | Ever |
| Other Respiratory Disease | 0/1 | ICD9 Code 277.0*<br>ICD9 Code 494*<br>ICD9 Code 515 | Ever |
| Hypertension | 0/1 | ICD9 Code 401*<br>ICD9 Code 402*<br>ICD9 Code 403*<br>ICD9 Code 404*<br>ICD9 Code 405* | Ever |
| Immunocompromised State | 0/1 | **Any of:**<br>ICD9 Code 042*<br>ICD9 Code 043*<br>ICD9 Code 044*<br>ICD9 Code<br>795.71 ICD9<br>Code V08 ICD9<br>Code V42.8*<br>ICD9 Proc Code 41.0*<br><br>**Or at least 2 of:** | For diagnosis codes, Ever<br>For drugs, 4 or more dispensed in last 12 months |

| | | |
|---|---|---|
| | | ATC4 Code<br>H02ABATC4 Code<br>H02BXATC4 Code<br>M01BA<br><br>**Or at least 2 of:**<br>ATC2 Code L04 |

**eTable 3: Results of Schoenfeld's Global test for proportional hazards assumption**

| | Model includes all ages | | | Model includes individuals ≥60 years of age | | |
|---|---|---|---|---|---|---|
| **Variable** | **Chisq** | **df** | **P** | **Chisq** | **df** | **p-Value** |
| Sex | 0.36 | 1 | 0.55 | 0.0000455 | 1 | 0.99 |
| Age | 1804.67 | 1 | <0.001 | - | - | - |
| Morning vs Evening | 138.27 | 1 | <0.001 | 10.2 | 1 | 0.001 |
| Diabetes | 515.29 | 1 | <0.001 | 2.46 | 1 | 0.12 |
| Obesity | 396.19 | 1 | <0.001 | 5.77 | 1 | 0.02 |
| Chronic kidney disease | 212.85 | 1 | <0.001 | 0.87 | 1 | 0.35 |
| Hypertension | 1478.49 | 1 | <0.001 | 0.98 | 1 | 0.32 |
| Global test | 2608.96 | 7 | <0.001 | 17.3 | 6 | 0.01 |

**eTable 4: Multivariate logistic regression model for association with COVID breakthrough infection post vaccine doses 1 and 2.**

| Variable | Odds Ratio | Confidence Interval (5%-95%) | p-Value |
|---|---|---|---|
| Third vaccine (booster) | 0.80 | 0.79-0.82 | <0.001 |
| Male gender | 0.81 | 0.80-0.82 | <0.001 |
| Evening doses | 1.02 | 1.01-1.04 | 0.006 |
| Diabetes Mellitus | 0.97 | 0.93-1.00 | 0.07 |
| Age > 60y | 0.68 | 0.66-0.70 | <0.001 |
| Obesity | 1.00 | 0.98-1.02 | 0.6 |
| Chronic Kidney Disease | 0.90 | 0.85-0.95 | <0.001 |
| Hypertension | 1.02 | 0.99-1.05 | 0.14 |

**eTable 5: Bootstrap multivariate analysis (2000 rounds) for the association with COVID infection post two vaccine doses.**

| Variable | Odds Ratio | Confidence Interval (5%-95%) |
|---|---|---|
| Third vaccine (booster) | 0.28 | 0.27-0.30 |
| Male gender | 0.82 | 0.80-0.83 |
| Evening doses | 1.07 | 1.06-1.09 |
| Diabetes Mellitus | 0.96 | 0.92-0.99 |
| Age > 60y | 0.52 | 0.50-0.55 |
| Obesity | 0.91 | 0.90-0.93 |
| Chronic Kidney Disease | 0.90 | 0.86-0.95 |
| Hypertension | 0.95 | 0.92-0.97 |

**eTable 6: Cox multivariate regression analysis of breakthrough infection after COVID-19 vaccination: morning vs. evening.**

| | Dose 1 and 2 | | | Dose 3 | | | Dose 4 | | |
|---|---|---|---|---|---|---|---|---|---|
| | HR | 95% CI | p | HR | 95% CI | p | HR | 95% CI | p |
| **Booster Dose**[a] | | | | | | | | | |
| **-** | — | — | | — | — | | — | — | |
| **+** | 0.48 | 0.48-0.49 | <0.001 | 0.44 | 0.42-0.45 | <0.001 | — | — | |
| **Vaccination Timing** | | | | | | | | | |
| **8:00-11:59** | 0.93 | 0.92-0.94 | <0.001 | 0.97 | 0.95-0.98 | <0.001 | 0.97 | 0.93-1.02 | 0.3 |
| **16:00-19:59** | — | — | | — | — | | — | — | — |
| **Sex** | | | | | | | | | |
| **F** | — | — | | — | — | | — | — | |
| **M** | 0.83 | 0.82-0.84 | <0.001 | 0.83 | 0.82-0.84 | <0.001 | 1.13 | 1.08-1.19 | <0.001 |
| **Age**[e] | | | | | | | | | |
| **≤60** | — | — | | — | — | | — | — | |
| **>60** | 0.66 | 0.64-0.67 | <0.001 | 0.68 | 0.67-0.70 | <0.001 | 1.17 | 1.10-1.24 | <0.001 |
| **Diabetes** | | | | | | | | | |
| **-** | — | — | | — | — | | — | — | |
| **+** | 0.92 | 0.89-0.96 | <0.001 | 0.91 | 0.89-0.94 | <0.001 | 0.89 | 0.84-0.94 | <0.001 |
| **BMI**[b,e] | | | | | | | | | |
| **<30** | — | — | | — | — | | — | — | |
| **≥30** | 0.91 | 0.90-0.93 | <0.001 | 0.94 | 0.93-0.95 | <0.001 | 0.95 | 0.90-1.00 | 0.052 |
| **CKD**[c] | | | | | | | | | |
| **-** | — | — | | — | — | | — | — | |
| **+** | 0.9 | 0.86-0.95 | <0.001 | 0.91 | 0.88-0.94 | <0.001 | 0.84 | 0.78-0.89 | <0.001 |
| **HTN**[d] | | | | | | | | | |
| **-** | — | — | | — | — | | — | — | |
| **+** | 0.93 | 0.91-0.96 | <0.001 | 0.93 | 0.91-0.95 | <0.001 | 0.93 | 0.88-0.98 | 0.004 |

[a]Models the effect one additional booster dose beyond the number of vaccinations specified.

[b]BMI, body mass index; [c]CKD, chronic kidney disease; [d]HTN, hypertension. [e]Modeling BMI and age as continuous variables yielded similar trends (data not shown).

**eTable 7: Cox multivariate regression analysis of breakthrough infection after COVID-19 vaccination: morning vs. afternoon administration.**

| | Dose 1 and 2 | | | Dose 3 | | | Dose 4 | | |
|---|---|---|---|---|---|---|---|---|---|
| | HR | 95% CI | p | HR | 95% CI | p | HR | 95% CI | p |
| **Booster Dose[a]** | | | | | | | | | |
| **-** | — | — | | — | — | | — | — | |
| **+** | 0.51 | 0.50-0.52 | <0.001 | 0.43 | 0.42-0.44 | <0.001 | — | — | |
| **Vaccination Timing** | | | | | | | | | |
| **8:00-11:59** | — | — | | — | — | | — | — | |
| **12:00-15:59** | 0.96 | 0.95-0.98 | <0.001 | 0.96 | 0.95-0.97 | <0.001 | 0.95 | 0.91-1 | 0.04 |
| **Sex** | | | | | | | | | |
| **F** | — | — | | — | — | | — | — | |
| **M** | 0.82 | 0.81-0.84 | <0.001 | 0.83 | 0.82-0.84 | <0.001 | 1.12 | 1.07-1.18 | <0.001 |
| **Age** | | | | | | | | | |
| **≤60** | — | — | | — | — | | — | — | |
| **>60** | 0.66 | 0.64-0.67 | <0.001 | 0.68 | 0.67-0.69 | <0.001 | 1.05 | 0.98-1.13 | 0.13 |
| **Diabetes** | | | | | | | | | |
| **-** | — | — | | — | — | | — | — | |
| **+** | 0.91 | 0.88-0.94 | <0.001 | 0.9 | 0.88-0.93 | <0.001 | 0.93 | 0.88-0.99 | 0.015 |
| **BMI[b,e]** | | | | | | | | | |
| **<30** | — | — | | — | — | | — | — | |
| **≥30** | 0.94 | 0.93-0.96 | <0.001 | 0.95 | 0.94-0.97 | <0.001 | 0.9 | 0.85-0.95 | <0.001 |
| **CKD[c]** | | | | | | | | | |
| **-** | — | — | | — | — | | — | — | |
| **+** | 0.92 | 0.88-0.97 | <0.001 | 0.9 | 0.87-0.93 | <0.001 | 0.83 | 0.78-0.89 | <0.001 |
| **HTN[d]** | | | | | | | | | |
| **-** | — | — | | — | — | | — | — | |
| **+** | 0.92 | 0.9-0.95 | <0.001 | 0.93 | 0.91-0.94 | <0.001 | 0.93 | 0.89-0.98 | 0.007 |

[a]Models the effect one additional booster dose beyond the number of vaccinations specified.

[b]BMI, body mass index; [c]CKD, chronic kidney disease; [d]HTN, hypertension. [e]Modeling BMI and age as continuous variables yielded similar trends (data not shown).

**eTable 8: Cox multivariate regression analysis of breakthrough infection after COVID-19 vaccination: afternoon vs. evening administration.**

| | Dose 1 and 2 | | | Dose 3 | | | Dose 4 | | |
|---|---|---|---|---|---|---|---|---|---|
| | **HR** | **95% CI** | **p** | **HR** | **95% CI** | **p** | **HR** | **95% CI** | **p** |
| **Booster Dose[a]** | | | | | | | | | |
| **-** | — | — | | — | — | | — | — | |
| **+** | 0.49 | 0.48-0.49 | <0.001 | 0.43 | 0.42-0.45 | <0.001 | — | — | |
| **Vaccination Timing** | | | | | | | | | |
| **12:00-15:59** | 0.88 | 0.87-0.90 | <0.001 | 0.93 | 0.91-0.94 | <0.001 | 0.93 | 0.88-0.97 | 0.003 |
| **16:00-19:59** | — | — | | — | — | | — | — | |
| **Sex** | | | | | | | | | |
| **F** | — | — | | — | — | | — | — | |
| **M** | 0.8 | 0.79-0.82 | <0.001 | 0.83 | 0.82-0.84 | <0.001 | 1.1 | 1.05-1.15 | <0.001 |
| **Age[e]** | | | | | | | | | |
| **≤60** | — | — | | — | — | | — | — | |
| **>60** | 0.68 | 0.66-0.7 | <0.001 | 0.69 | 0.67-0.70 | <0.001 | 1.1 | 1.03-1.17 | 0.003 |
| **Diabetes** | | | | | | | | | |
| **-** | — | — | | — | — | | — | — | |
| **+** | 0.94 | 0.9-0.98 | 0.002 | 0.93 | 0.92-0.95 | <0.001 | 0.92 | 0.87-0.97 | 0.003 |
| **BMI[b,e]** | | | | | | | | | |
| **<30** | — | — | | — | — | | — | — | |
| **≥30** | 0.9 | 0.89-0.92 | <0.001 | 0.93 | 0.92-0.95 | <0.001 | 0.95 | 0.90-1.00 | 0.052 |
| **CKD[3]** | | | | | | | | | |
| **-** | — | — | | — | — | | — | — | |
| **+** | 0.94 | 0.89-0.99 | <0.001 | 0.89 | 0.86-0.93 | <0.001 | 0.83 | 0.78-0.89 | <0.001 |
| **HTN[d]** | | | | | | | | | |
| **-** | — | — | | — | — | | — | — | |
| **+** | 0.94 | 0.91-0.97 | <0.001 | 0.95 | 0.93-0.97 | <0.001 | 0.94 | 0.89-0.99 | 0.013 |

[a]Models the effect one additional booster dose beyond the number of vaccinations specified.

[b]BMI, body mass index; [c]CKD, chronic kidney disease; [d]HTN, hypertension. [e]Modeling BMI and age as continuous variables yielded similar trends (data not shown).

**eTable 9: Cox multivariate regression analysis of emergency department (ED) visits after COVID-19 vaccination.**

| | Dose 1 and 2[a] HR | Dose 1 and 2[a] 95% CI | Dose 1 and 2[a] p | Dose 3 HR | Dose 3 95% CI | Dose 3 p |
|---|---|---|---|---|---|---|
| **Booster Dose[b]** | | | | | | |
| **-** | — | — | | — | — | |
| **+** | 0.26 | 0.22-0.30 | <0.001 | 0.33 | 0.27-0.41 | <0.001 |
| **Vaccination Timing** | | | | | | |
| **8:00-11:59** | 0.99 | 0.83-1.13 | >0.9 | 1.04 | 0.92-1.17 | 0.5 |
| **16:00-19:59** | — | — | | — | — | — |
| **Age[f]** | | | | | | |
| **≤60** | — | — | | — | — | |
| **>60** | 2.88 | 2.37-3.50 | <0.001 | 1.99 | 1.64-2.42 | <0.001 |
| **BMI[c,f]** | | | | | | |
| **≥30** | — | — | | — | — | |
| **<30** | 1.23 | 1.04-1.45 | 0.014 | 1.01 | 0.86-1.20 | 0.9 |
| **Diabetes** | | | | | | |
| **-** | — | — | | — | — | |
| **+** | 1.5 | 1.22-1.86 | <0.001 | 1.51 | 1.24-1.85 | <0.001 |
| **HTN[d]** | | | | | | |
| **-** | — | — | | — | — | |
| **+** | 1.23 | 1.00-1.52 | 0.046 | 1.36 | 1.11-1.65 | 0.002 |
| **CKD[e]** | | | | | | |
| **-** | — | — | | — | — | |
| **+** | 2.63 | 2.09-3.30 | <0.001 | 2.63 | 2.11-3.27 | <0.001 |
| **Immuno-suppression** | | | | | | |
| **-** | — | — | | — | — | |
| **+** | 4.77 | 3.83-5.94 | <0.001 | 5.21 | 4.25-6.40 | <0.001 |

[a]Both doses were administered within the indicated time frame. [b]Models the effect one additional booster dose beyond the number of vaccinations specified. [c]Body Mass Index. [d]Hypertension. [e]Chronic kidney disease. [f]Modeling BMI and age as continuous variables yielded similar trends (data not shown).

**eTable 10: Patient demographic breakdown for periodicity analysis of breakthrough infections (Fig. 3)**

| | Doses 1 and 2 | | | Dose 3 | | | Dose 4 | | |
|---|---|---|---|---|---|---|---|---|---|
| **Time Bin** | **Mean Age (SD)** | **Male (%)** | **≥ 4 Comorbidities** | **Mean Age (SD)** | **Male (%)** | **≥ 4 Comorbidities** | **Mean Age (SD)** | **Male (%)** | **≥ 4 Comorbidities** |
| **8:00-9:59** | 42.1 (17.3) | 49.0% | 1.0% | 47.3 (17.8) | 47.9% | 1.8% | 68.3 (11.2) | 52.5% | 5.7% |
| **10:00-11:59** | 40.9 (18.5) | 46.8% | 1.3% | 46.4 (18.6) | 46.9% | 1.9% | 69.8 (11.1) | 49.1% | 6.4% |
| **11:00-12:59** | 40.4 (18.3) | 47.3% | 1.2% | 46.3 (18.6) | 47.4% | 1.8% | 70.3 (10.8) | 48.6% | 6.4% |
| **12:00-13:59** | 40.0 (18.1) | 47.1% | 1.1% | 47.0 (18.7) | 47.8% | 1.9% | 70.6 (10.4) | 48.6% | 6.4% |
| **13:00-14:59** | 38.4 (18) | 46.7% | 0.9% | 47.2 (18.9) | 48.1% | 1.9% | 69.7 (11.2) | 48.7% | 6.2% |
| **14:00-15:59** | 37.2 (18.1) | 46.4% | 0.8% | 46.2 (19.2) | 47.8% | 1.9% | 68.5 (11.9) | 49.7% | 5.9% |
| **15:00-16:59** | 36.3 (18.2) | 45.7% | 0.7% | 44.7 (19.4) | 47.0% | 1.7% | 67.1 (12.3) | 50.6% | 5.6% |
| **16:00-17:59** | 35.6 (17.9) | 46.4% | 0.7% | 43.8 (19.3) | 47.6% | 1.5% | 65.6 (12.6) | 51.9% | 5.2% |
| **17:00-18:59** | 36.5 (17.3) | 48.6% | 0.6% | 43.5 (19.0) | 49.1% | 1.4% | 65.0 (12.3) | 54.4% | 4.8% |
| **18:00-19:59** | 38.2 (16) | 50.0% | 0.5% | 43.9 (18.7) | 50.3% | 1.4% | 65.7 (11.4) | 55.7% | 4.8% |
| **19:00-20:59** | 38.7 (15.5) | 50.2% | 0.5% | 45.0 (18.1) | 50.8% | 1.3% | 67.3 (10.2) | 55.3% | 5.3% |
| **20:00-20:59** | 37.6 (15.1) | 49.3% | 0.3% | 45.5 (17.4) | 51.9% | 1.3% | 68.4 (9.1) | 55.6% | 6.0% |

**eTable 11: Breakthrough COVID-19 infection hazard ratio (HR) comparing AM (8:00-9:59) vaccination to the indicated times. Ratios >1 indicate fewer infections associated with the AM reference time. For graphical representation of these data see Figure 3A-C.**

| Time Bin | Dose 1 and 2 HR | Dose 1 and 2 95% CI | Dose 1 and 2 n | Dose 3 HR | Dose 3 95% CI | Dose 3 n | Dose 4 HR | Dose 4 95% CI | Dose 4 n |
|---|---|---|---|---|---|---|---|---|---|
| 08:00-09:59 | 1.00 | | 98787 | 1.00 | | 186932 | 1.00 | | 29870 |
| 10:00-11:59 | 0.93 | 0.91-0.95 | 83523 | 0.98 | 0.96-0.99 | 204203 | 0.99 | 0.92-1.05 | 35345 |
| 11:00-12:59 | 0.92 | 0.90-0.94 | 75240 | 0.97 | 0.96-0.99 | 191679 | 0.96 | 0.89-1.02 | 32442 |
| 12:00-13:59 | 0.91 | 0.88-0.93 | 65926 | 0.94 | 0.93-0.96 | 181319 | 0.96 | 0.89-1.02 | 32471 |
| 13:00-14:59 | 0.94 | 0.92-0.96 | 65274 | 0.93 | 0.92-0.95 | 173612 | 0.90 | 0.84-0.96 | 36958 |
| 14:00-15:59 | 0.96 | 0.93-0.98 | 71112 | 0.96 | 0.95-0.98 | 170538 | 0.90 | 0.84-0.96 | 42222 |
| 15:00-16:59 | 1.01 | 0.98-1.03 | 80343 | 1.00 | 0.98-1.01 | 186127 | 0.94 | 0.89-1.01 | 47780 |
| 16:00-17:59 | 1.07 | 1.04-1.09 | 90676 | 1.02 | 1.00-1.03 | 206931 | 0.99 | 0.93-1.06 | 47576 |
| 17:00-18:59 | 1.06 | 1.03-1.08 | 85023 | 1.02 | 1.01-1.04 | 179406 | 1.04 | 0.98-1.11 | 35991 |
| 18:00-19:59 | 0.99 | 0.97-1.01 | 64325 | 1.03 | 1.01-1.05 | 114029 | 1.08 | 1.00-1.17 | 18538 |
| 19:00-20:59 | 0.96 | 0.93-0.99 | 29769 | 1.02 | 0.99-1.04 | 54557 | 1.16 | 1.05-1.29 | 6138 |
| 20:00-21:59 | 0.87 | 0.82-0.93 | 5887 | 1.01 | 0.97-1.05 | 16326 | 1.16 | 0.95-1.42 | 1458 |

**eTable 12: COVID-19 associated ED visit hazard ratio (HR) comparing AM (8:00-9:59) vaccination to the indicated times. Ratios >1 indicate fewer infections associated with the AM reference time. For graphical representation of these data see Figure 3D.**

| | Dose 1 and 2 | | | Dose 3 | | |
|---|---|---|---|---|---|---|
| **Time Bin** | **HR** | **95% CI** | **n** | **HR** | **95% CI** | **n** |
| **08:00-09:59** | 1.00 | | 98787 | 1.00 | | 186932 |
| **10:00-11:59** | 0.84 | 0.67-1.05 | 83523 | 0.88 | 0.73-1.05 | 204203 |
| **11:00-12:59** | 0.98 | 0.78-1.22 | 75240 | 0.90 | 0.75-1.08 | 191679 |
| **12:00-13:59** | 1.09 | 0.87-1.37 | 65926 | 0.88 | 0.73-1.07 | 181319 |
| **13:00-14:59** | 0.94 | 0.73-1.20 | 65274 | 0.82 | 0.68-1.00 | 173612 |
| **14:00-15:59** | 0.97 | 0.76-1.23 | 71112 | 0.79 | 0.64-0.96 | 170538 |
| **15:00-16:59** | 0.95 | 0.75-1.20 | 80343 | 0.95 | 0.79-1.15 | 186127 |
| **16:00-17:59** | 1.00 | 0.80-1.26 | 90676 | 0.89 | 0.74-1.07 | 206931 |
| **17:00-18:59** | 1.04 | 0.83-1.31 | 85023 | 0.80 | 0.66-0.98 | 179406 |
| **18:00-19:59** | 1.04 | 0.81-1.33 | 64325 | 0.91 | 0.73-1.14 | 114029 |
| **19:00-20:59** | 0.95 | 0.67-1.33 | 29769 | 1.10 | 0.84-1.44 | 54557 |
| **20:00-21:59** | 0.46 | 0.17-1.23 | 5887 | 1.28 | 0.84-1.95 | 16326 |

**eTable 13: METACYLE output for data presented in Figures 3 and 4.**

| COVID Vaccination Number | Age Range | Outcome | meta2d pvalue | meta2d BH.Q | meta2d period | meta2d phase | meta2d Base | meta2d AMP | meta2d rAMP |
|---|---|---|---|---|---|---|---|---|---|
| **Dose 1/2** | All | Breakthrough Infection | 0.001 | 0.004 | 14.018 | 12.525 | 0.948 | 0.062 | 0.062 |
| | 12-30 Years | Breakthrough Infection | 0.003 | 0.006 | 13.325 | 0.120 | 0.915 | 0.096 | 0.096 |
| | 30-60 Years | Breakthrough Infection | 0.006 | 0.010 | 14.650 | 12.324 | 0.951 | 0.028 | 0.028 |
| | >60 Years | Breakthrough Infection | 0.124 | 0.151 | 16.388 | 11.111 | 0.963 | 0.034 | 0.034 |
| | All | ED Visit | 0.153 | 0.153 | 11.466 | 5.402 | 0.928 | 0.105 | 0.105 |
| **Dose 3** | All | Breakthrough Infection | 0.003 | 0.006 | 14.823 | 11.964 | 0.993 | 0.017 | 0.017 |
| | 12-30 Years | Breakthrough Infection | <0.001 | 0.003 | 15.157 | 12.184 | 1.009 | 0.006 | 0.006 |
| | 30-60 Years | Breakthrough Infection | 0.020 | 0.028 | 13.554 | 13.532 | 0.966 | 0.011 | 0.011 |
| | >60 Years | Breakthrough Infection | 0.006 | 0.010 | 16.747 | 13.022 | 1.048 | 0.062 | 0.060 |
| | All | ED Visit | 0.152 | 0.153 | 14.055 | 10.113 | 0.928 | 0.022 | 0.022 |
| **Dose 4** | All | Breakthrough Infection | <0.001 | <0.001 | 16.077 | 13.563 | 1.049 | 0.105 | 0.100 |

**eTable 14: Breakthrough COVID-19 infection hazard ratio (HR) as a function of age bracket, comparing AM (8:00-11:59) to PM (16:00-19:59) vaccination times. Ratios >1 indicate fewer infections associated with the AM reference time. For graphical representation of these data see Figure 4A.**

| Age Bin [Years] | Dose 1 and 2 | | | Dose 3 | | |
|---|---|---|---|---|---|---|
| | **HR** | **95% CI** | **Total n** | **HR** | **95% CI** | **Total n** |
| **12-30** | 1.07 | 1.04-1.09 | 156845 | 1.10 | 1.08-1.13 | 175070 |
| **30-60** | 1.03 | 1.01-1.05 | 246832 | 0.98 | 0.96-0.99 | 368111 |
| **>60** | 1.12 | 1.07-1.18 | 60370 | 1.07 | 1.03-1.10 | 168860 |

**eTable 15: Breakthrough COVID-19 infection hazard ratio (HR) in patients 12-30 years old, comparing AM (8:00-9:59) vaccination to the indicated times. Ratios >1 indicate fewer infections associated with the AM reference time. For graphical representation of these data see Figure 4B.**

| | Dose 1 and 2 | | | Dose 3 | | |
|---|---|---|---|---|---|---|
| **Time Bin** | **HR** | **95% CI** | **n** | **HR** | **95% CI** | **n** |
| **08:00-09:59** | 1.00 | | 25502 | 1.00 | | 35445 |
| **10:00-11:59** | 0.94 | 0.90-0.97 | 27084 | 0.97 | 0.93-1.00 | 47677 |
| **11:00-12:59** | 0.90 | 0.87-0.94 | 25632 | 0.96 | 0.93-0.99 | 45992 |
| **12:00-13:59** | 0.88 | 0.85-0.92 | 22992 | 0.92 | 0.89-0.96 | 41710 |
| **13:00-14:59** | 0.90 | 0.86-0.93 | 24923 | 0.92 | 0.89-0.96 | 39803 |
| **14:00-15:59** | 0.90 | 0.86-0.94 | 29103 | 0.98 | 0.95-1.01 | 42315 |
| **15:00-16:59** | 0.98 | 0.94-1.02 | 34537 | 1.04 | 1.00-1.07 | 51461 |
| **16:00-17:59** | 1.08 | 1.05-1.13 | 39777 | 1.07 | 1.04-1.11 | 59998 |
| **17:00-18:59** | 1.05 | 1.01-1.09 | 34995 | 1.09 | 1.05-1.13 | 52446 |
| **18:00-19:59** | 0.94 | 0.91-0.98 | 23416 | 1.10 | 1.06-1.14 | 31950 |
| **19:00-20:59** | 0.88 | 0.83-0.92 | 10075 | 1.06 | 1.01-1.11 | 13738 |
| **20:00-21:59** | 0.80 | 0.73-0.89 | 2107 | 1.00 | 0.93-1.09 | 3810 |

**eTable 16: Breakthrough COVID-19 infection hazard ratio (HR) in patients 30-60 years old, comparing AM (8:00-9:59) vaccination to the indicated times. Ratios >1 indicate fewer infections associated with the AM reference time. For graphical representation of these data see Figure 4C.**

| | Dose 1 and 2 | | | Dose 3 | | |
|---|---|---|---|---|---|---|
| **Time Bin** | **HR** | **95% CI** | **n** | **HR** | **95% CI** | **n** |
| **08:00-09:59** | 1.00 | | 58496 | 1.00 | | 103783 |
| **10:00-11:59** | 0.93 | 0.91-0.96 | 42817 | 0.99 | 0.97-1.01 | 105491 |
| **11:00-12:59** | 0.92 | 0.90-0.95 | 38015 | 0.97 | 0.95-0.99 | 98468 |
| **12:00-13:59** | 0.91 | 0.88-0.94 | 33272 | 0.95 | 0.93-0.97 | 92407 |
| **13:00-14:59** | 0.94 | 0.91-0.97 | 31863 | 0.93 | 0.91-0.95 | 86538 |
| **14:00-15:59** | 0.95 | 0.93-0.98 | 33655 | 0.95 | 0.93-0.97 | 83818 |
| **15:00-16:59** | 0.98 | 0.95-1.01 | 37148 | 0.96 | 0.94-0.98 | 90551 |
| **16:00-17:59** | 1.00 | 0.97-1.03 | 42052 | 0.97 | 0.95-0.99 | 101528 |
| **17:00-18:59** | 1.01 | 0.98-1.04 | 41927 | 0.97 | 0.95-0.99 | 88891 |
| **18:00-19:59** | 0.99 | 0.96-1.02 | 34695 | 0.98 | 0.95-1.00 | 57329 |
| **19:00-20:59** | 1.00 | 0.96-1.04 | 17022 | 0.97 | 0.94-1.00 | 28452 |
| **20:00-21:59** | 0.91 | 0.84-0.98 | 3346 | 0.97 | 0.92-1.02 | 8778 |

**eTable 17: Breakthrough COVID-19 infection hazard ratio (HR) in patients >60 years old, comparing AM (8:00-9:59) vaccination to the indicated times. Ratios >1 indicate fewer infections associated with the AM reference time. For graphical representation of these data see Figure 4C.**

| | Dose 1 and 2 | | | Dose 3 | | |
|---|---|---|---|---|---|---|
| **Time Bin** | **HR** | **95% CI** | **n** | **HR** | **95% CI** | **n** |
| **08:00-09:59** | 1.00 | | 14789 | 1.00 | | 47704 |
| **10:00-11:59** | 0.84 | 0.78-0.90 | 13622 | 0.97 | 0.93-1.01 | 51035 |
| **11:00-12:59** | 0.89 | 0.83-0.96 | 11593 | 0.99 | 0.95-1.03 | 47219 |
| **12:00-13:59** | 0.91 | 0.85-0.98 | 9662 | 0.96 | 0.92-1.00 | 47202 |
| **13:00-14:59** | 0.94 | 0.86-1.01 | 8488 | 0.94 | 0.90-0.98 | 47271 |
| **14:00-15:59** | 1.00 | 0.92-1.08 | 8354 | 0.97 | 0.93-1.01 | 44405 |
| **15:00-16:59** | 0.95 | 0.88-1.03 | 8658 | 1.01 | 0.97-1.05 | 44115 |
| **16:00-17:59** | 0.97 | 0.90-1.05 | 8847 | 1.04 | 1.00-1.09 | 45405 |
| **17:00-18:59** | 1.07 | 0.99-1.16 | 8101 | 1.05 | 1.01-1.10 | 38069 |
| **18:00-19:59** | 1.07 | 0.99-1.16 | 6214 | 1.06 | 1.01-1.12 | 24750 |
| **19:00-20:59** | 1.03 | 0.92-1.16 | 2672 | 1.09 | 1.02-1.16 | 12367 |
| **20:00-21:59** | 0.98 | 0.75-1.28 | 434 | 1.19 | 1.08-1.32 | 3738 |

**eTable 18: Distribution of COVID-19 tests in the study population. See eFigure 2 for a graphical presentation of these data.**

| Hour Bin | Total Tests | | Positive Tests | | Positivity Rate | |
|---|---|---|---|---|---|---|
| | PM[a] | AM[a] | PM | AM | PM | AM |
| **0** | 18301 | 27141 | 509 | 779 | 0.028 | 0.029 |
| **1** | 6218 | 8880 | 52 | 92 | 0.008 | 0.010 |
| **2** | 5496 | 7990 | 33 | 52 | 0.006 | 0.007 |
| **3** | 6212 | 8998 | 48 | 63 | 0.008 | 0.007 |
| **4** | 3776 | 5464 | 50 | 75 | 0.013 | 0.014 |
| **5** | 1951 | 2553 | 42 | 58 | 0.022 | 0.023 |
| **6** | 9333 | 13285 | 55 | 101 | 0.006 | 0.008 |
| **7** | 7562 | 11183 | 255 | 337 | 0.034 | 0.030 |
| **8** | 23481 | 35000 | 1970 | 2552 | 0.084 | 0.073 |
| **9** | 36674 | 55411 | 3518 | 4766 | 0.096 | 0.086 |
| **10** | 41152 | 61818 | 4267 | 5804 | 0.104 | 0.094 |
| **11** | 42229 | 60857 | 4240 | 5895 | 0.100 | 0.097 |
| **12** | 46097 | 64411 | 4879 | 6246 | 0.106 | 0.097 |
| **13** | 39846 | 52965 | 3982 | 5017 | 0.100 | 0.095 |
| **14** | 35845 | 45797 | 3448 | 4197 | 0.096 | 0.092 |
| **15** | 34279 | 42387 | 2993 | 3469 | 0.087 | 0.082 |
| **16** | 35709 | 40957 | 2903 | 3145 | 0.081 | 0.077 |
| **17** | 38150 | 41949 | 2894 | 2900 | 0.076 | 0.069 |
| **18** | 32168 | 34999 | 2632 | 2691 | 0.082 | 0.077 |
| **19** | 25905 | 28451 | 2260 | 2274 | 0.087 | 0.080 |
| **20** | 19160 | 23005 | 1405 | 1488 | 0.073 | 0.065 |
| **21** | 15003 | 18950 | 945 | 1036 | 0.063 | 0.055 |
| **22** | 9372 | 12952 | 291 | 322 | 0.031 | 0.025 |
| **23** | 8142 | 11692 | 99 | 136 | 0.012 | 0.012 |

[a]AM:8:00-11:59; PM:16:00-19:59
